# The Efficacy of Contact Tracing for the Containment of the 2019 Novel Coronavirus (COVID-19)

**DOI:** 10.1101/2020.02.14.20023036

**Authors:** Matt J Keeling, T Déirdre Hollingsworth, Jonathan M Read

## Abstract

Contact tracing is a central public health response to infectious disease outbreaks, especially in the early stages of an outbreak when specific treatments are limited. Importation of novel Coronavirus (COVID-19) from China and elsewhere into the United Kingdom highlights the need to understand the impact of contact tracing as a control measure. Using detailed survey information on social encounters coupled to predictive models, we investigate the likely efficacy of the current UK definition of a close contact (within 2 meters for 15 minutes or more) and the distribution of secondary cases that may go untraced. Taking recent estimates for COVID-19 transmission, we show that less than 1 in 5 cases will generate any subsequent untraced cases, although this comes at a high logistical burden with an average of 36.1 individuals (95^th^ percentiles 0-182) traced per case. Changes to the definition of a close contact can reduce this burden, but with increased risk of untraced cases; we estimate that any definition where close contact requires more than 4 hours of contact is likely to lead to uncontrolled spread.

---

Contact tracing is the main public health response to importations of rare or emerging infectious diseases, and was implemented in the UK during the ‘containment stage’ of the 2009 influenza pandemic (McLean et al 2010). In more recent years, contact tracing was also a valuable tool following importation of Ebola virus disease into the UK in 2014 (Crook et al 2017) and the cases of monkeypox in the UK in 2018 (Vaughan et al 2018). In general, contact tracing is a highly effective and robust strategy given sufficient resources. The main advantages are that it can identify potentially infected individuals before severe symptoms emerge, and if conducted sufficiently quickly can prevent onward transmission from the secondary cases. Contact tracing has proved hugely successful in the treatment of sexually transmitted infections, where the definition of a contact is relatively straightforward, where infection is often asymptomatic and where the time-scales of transmission are slow (Hogben et al 2016, Rönn et al 2017). In contrast, the use of contact tracing for novel invading pathogens has received less quantitative consideration, in part due to greater uncertainties over social contact structure (although see Ahmed et al 2018, Hoang et al 2019). Modelling studies have often focused on quantifying the importance of pre-symptomatic and pre-tracing infectiousness, but are usually based on statistical distributions of contact networks (Fraser et al 2004, Kwok et al 2019). Here we leverage detailed social network data from the UK to model both transmission and the act of tracing, and identify the implications of contact tracing for containment of a novel pathogen, using parameters for the novel Coronavirus (COVID-19) (Read et al 2020, Li et al 2020).

We characterised contact patterns in the UK using a postal and online cross-sectional survey, which asked participants to report the number of social encounters with unique individuals during a given day, as well as the duration and typical frequency of those encounters (Danon et al 2012, 2013). In total, 5,802 respondents reported more than 50,000 encounters - one of the biggest studies of its kind to date. The encounter patterns of this study were in good qualitative agreement with other similar studies of social interactions (Mossong et al 2008, Isella et al 2010). In this study, the daily encounter data was first extrapolated to generate a pattern of contacts over a 14 day period (replicating random encounters and increasing the total duration associated regular contacts), to act as the basis for transmission and contact tracing simulations. Using this extrapolated data, we can classify interactions into those which satisfy the definition of a close contact for the purpose of contact tracing. From our social encounter data we can also distinguish interactions with people who could be later identified and traced, from those with unidentifiable strangers (schematic figure 1). We assume that all contact of longer than 1 hour or repeated contacts can be identified and traced, whereas shorter meetings with people for the first time are strangers who are unidentifiable. The second element of the simulation is to determine who gets infected from a source case chosen representatively from the survey respondents. This transmission process is stochastic, accounting for both the time spent with each contact and the infectivity on each day (see Appendix). Taken together these two predictions allow us to bound the efficacy of contact tracing.

**FIGURE 1.**
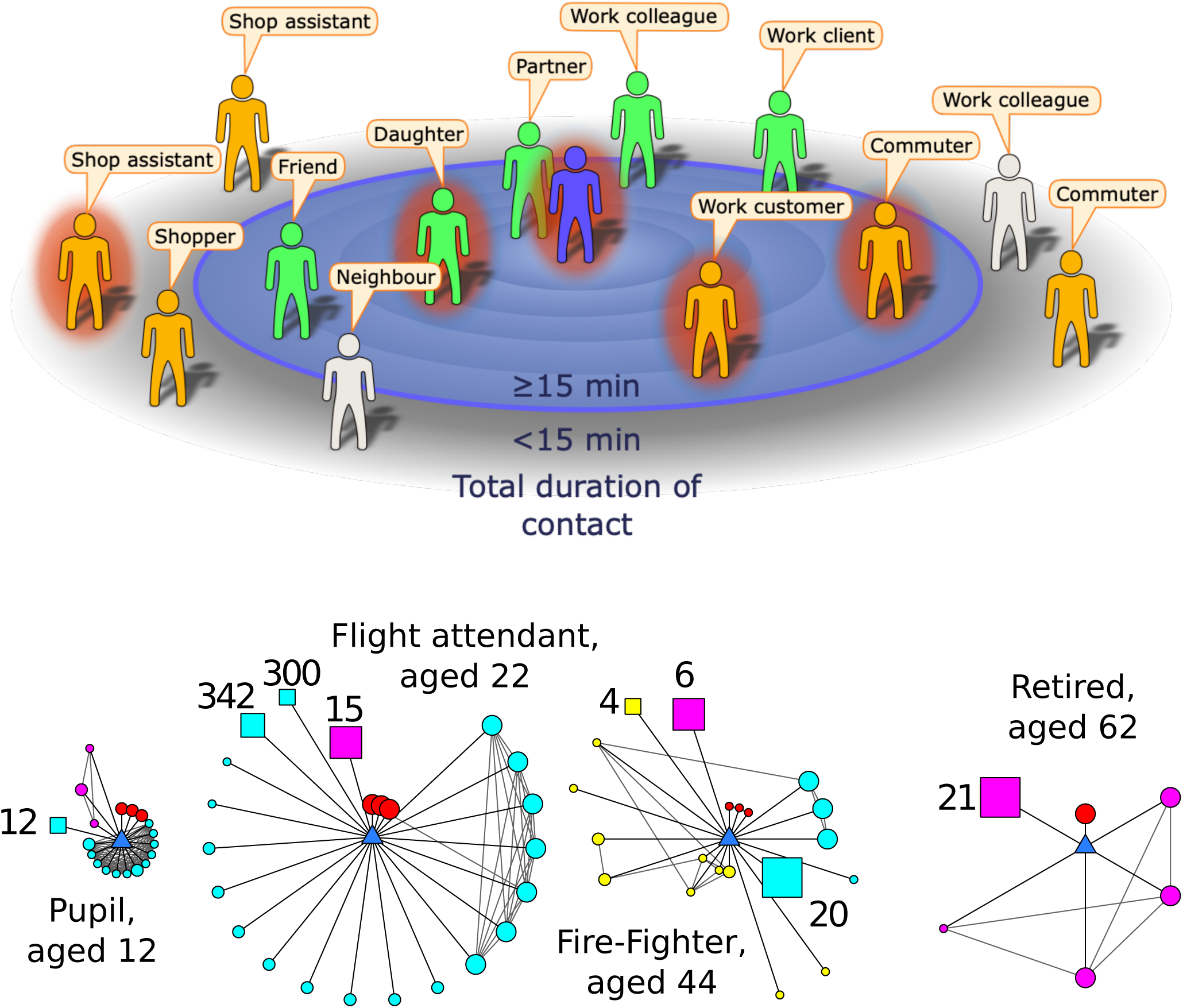
a) Cartoon example of the encounters made during a day by an infectious index case (central figure) with contacts positioned by their total contact duration. Here, the definition of a contact is someone with whom the index case encountered for 15 minutes or longer. Some contacts will be identifiable (green), while others will be unidentifiable (orange). A definition of contact that is too restrictive and inappropriate for the infection means some encounters may fail to meet the definition yet may be at risk of infection; these excluded contacts could be identifiable (light grey) or unidentifiable (orange). (b) Examples of ego-centric networks collected by the survey. The participant (ego) is the blue central triangle; circles represent individual contacts, squares represent groups of contacts (size of group indicated). Colours represent social settings of encounters (red=home, cyan=work/school, yellow=travel, pink=other). Larger symbol sizes represent longer contact durations, while a closer proximity to the ego indicates the contact is more frequently encountered.

## Heterogeneity in Behaviour

One of the most notable features of human social contacts is the huge variability in the number and strength of contacts - which is reflected as variation in both the number of secondary cases and the number of individuals that match the contact-tracing definition (figure 2). Using preliminary estimates of COVID-19 transmission (average latent period 4 days, average effective infectious period 1.61 days, *R*_0_=3.11 and assuming a simple SEIR formulation (Read et al 2020)) we compute the distribution of epidemiological, social and contact tracing characteristics across the population. Extrapolating the data from the social contact survey suggests that the average number of contacts over a 14 day period is 217, although the distribution is significantly over dispersed (with a median of 90 and around 3% of individuals having >1,000 total contacts). Of these total encounters, an average of 59 contacts (27%) meet the definition of a close-contact (in contact for >15 minutes, PHE 2020) and of these close-contacts we predict an average of 36 (61%) to be individuals known to the infected case that can be traced. Therefore, simply considering social contacts, it is clear that there are very many short duration contacts which do not meet the definition of a close contact, and although unlikely to become infected may pose a risk due to their greater abundance.

**FIGURE 2.**
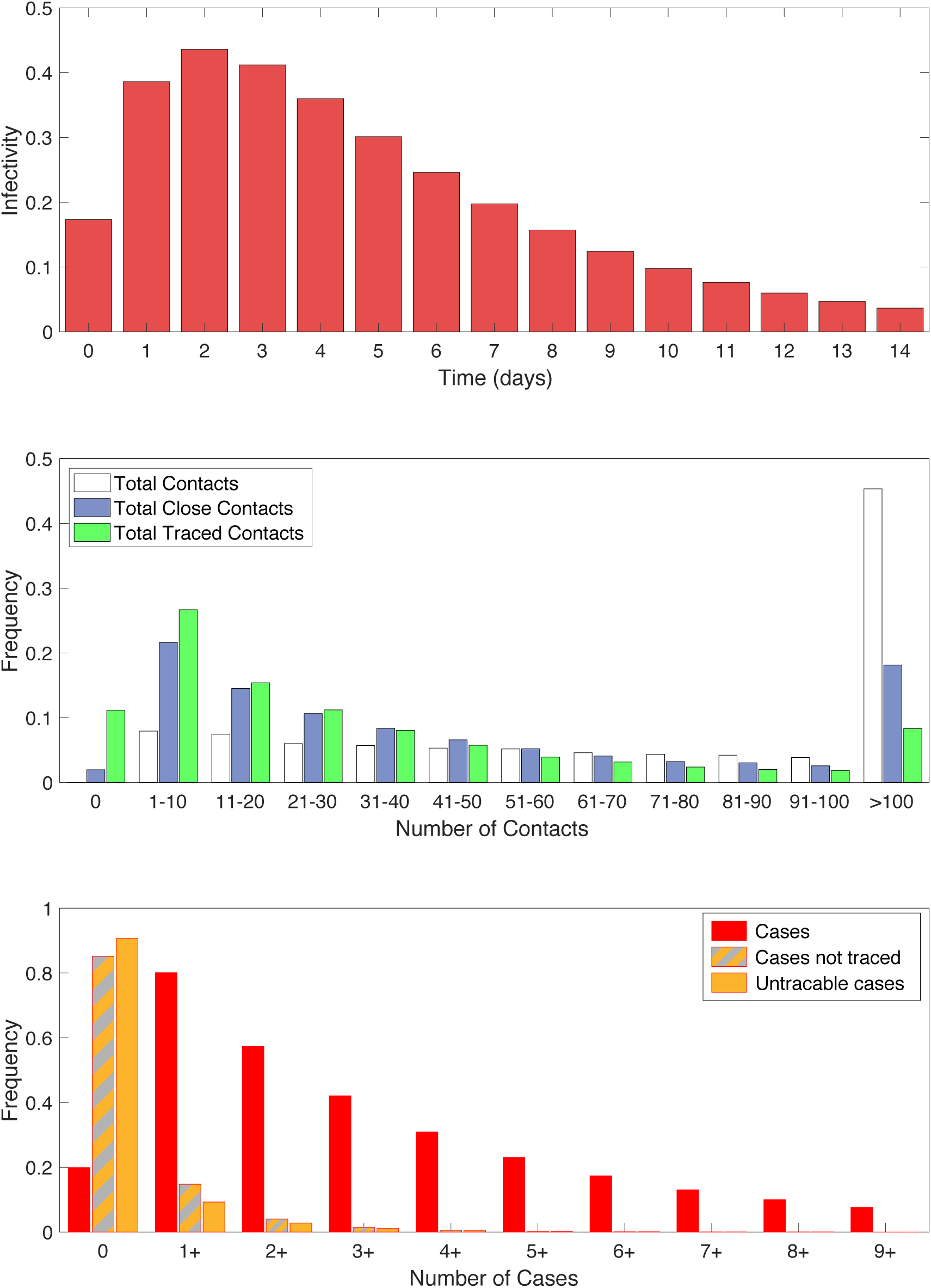
Distributions associated with transmission and contact tracing. (a) Relative infectivity over time based on an SEIR model with latent period 4, infectious period 1.61, *R*_0_=3.11. (b) Frequency distribution of the number contacts using colours from Figure 1a: white is all contacts; blue are those matching the >15 minute definition of a close contact; green are those matching the definition that are also identifiable. (c) Frequency distribution of the number of cases, again using colours from Figure 1a: red is all secondary cases; grey and orange are those that are not traced either through failing to meet the definition or because they are unidentified; orange are all secondary cases that are unidentifiable.

Given that the risk of infection increases with duration of contact, the distribution of cases effectively represents a biased sample of all contacts. As expected, given the model assumptions, the expected number of total secondary cases agrees with the assumed *R*_0_ (mean=3.11, median=2, and 95th percentiles 0-10). Given that these cases are most likely to be those contacts of the longest duration, we predict that 95% of cases match the definition of a close contact. However, not all of these contacts will be identifiable; assuming that all repeated contacts and contact of longer than 1 hour can be traced, we predict that 93% of all cases meet the definition and can be identified. However, because of the extreme heterogeneity in contacts between individuals and the stochastic nature of transmission, we would still expect 15% of all primary cases to generate at least one secondary case that cannot be identified. Aggregating across all individuals and under the optimistic assumption that all the contact tracing can be performed rapidly, we expect contact tracing to reduce the basic reproductive ratio from 3.11 to 0.21 - enabling the outbreak to be contained (figure 2).

Rapid and effective contact tracing can therefore be highly effective in the early control of COVID-19, but places substantial demands on the local public-health authorities. Each new case requires an average of 36 individuals to be traced, with 8.7% of cases having more than 100 close traceable contacts (figure 2). We therefore consider the implications of changing the definition of a close contact. Clearly a more strict definition of a close contact (requiring more contact time) reduces the burden on the health services as fewer contacts need to be traced, but also increases the risk of cases being missed. Figure 3 provides a quantitative assessment of changes to the close contact definition. Definitions requiring more than four hours of contact, are unlikely to control an outbreak as the expected number of untraced secondly cases is greater than one. This therefore places a strict bound on level of contact tracing required. The added benefit from definitions shorter than 1 hour has relatively little impact on the mean number of untraced cases (figure 3b), but does reduce the probability that some untraced contacts occur.

**FIGURE 3.**
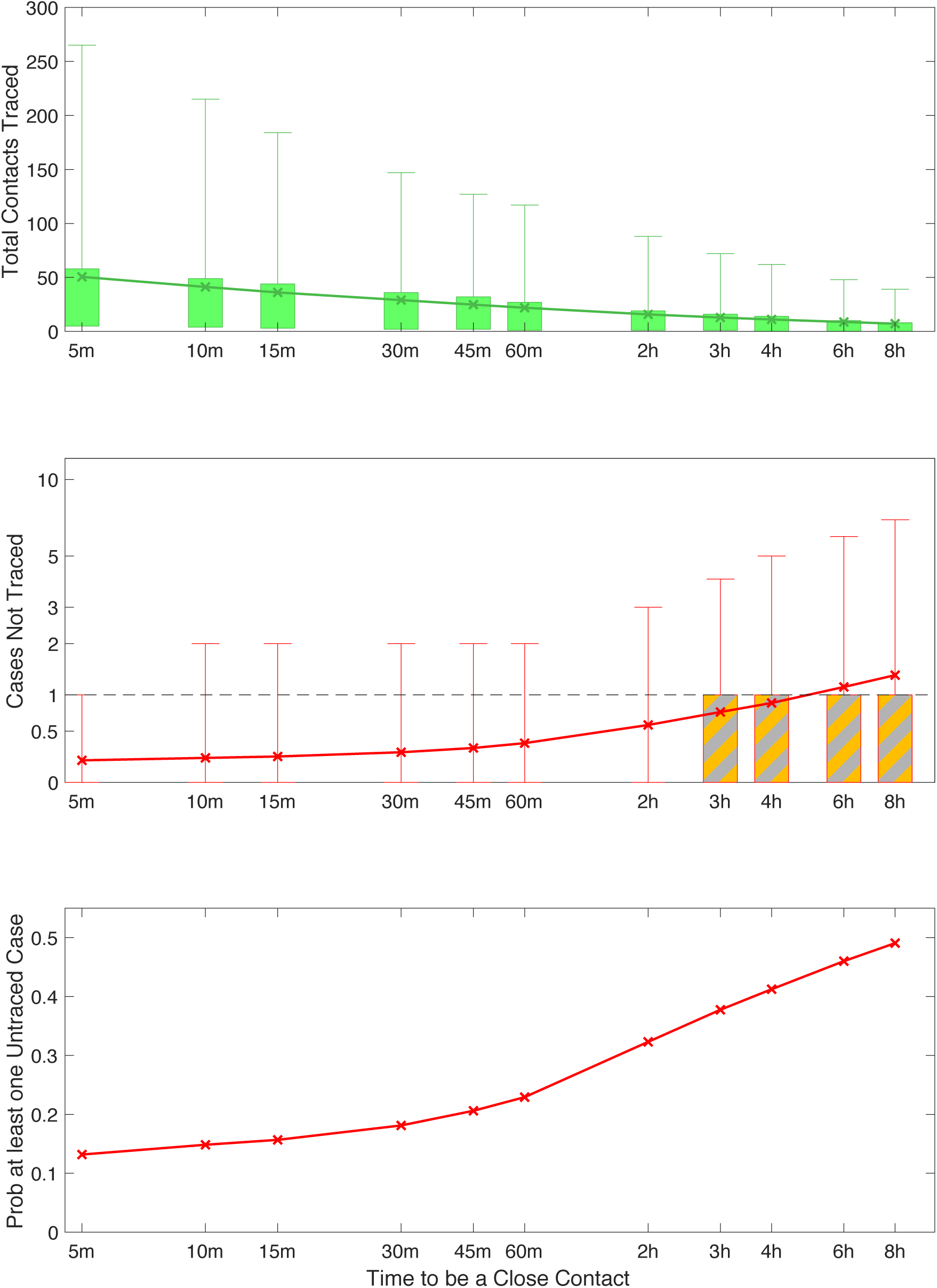
Impact of different assumptions for the definition of a close-contact. (a) the total number of contacts traced (b) the number of secondary contacts that are not traced (c) the probability that at least one secondary case is not traced. For (a) and (b) the crosses mark the mean value, boxes contain the 50th percentiles while bars contact the 95th percentiles, colours correspond to those in Figure 1a - distributions are across all respondents to the survey and across stochastic realisations. (Based on an SEIR model with latent period 4, infectious period 1.61, *R*_0_=3.11).

Throughout we have used a value of *R*_0_ that represents a population-level average once the local infection has become established. However, the first invasion into any new population or social setting generally has a larger expected number of secondary cases. The first invader enters a completely susceptible population; moreover all their close contacts (eg family members) are susceptible. In contrast, due to clustering of contacts, most secondary cases will be in a landscape with a depleted number of susceptibles - as close contacts such as family members will already have been exposed to the primary case. This susceptible depletion in the local social network may help to explain the change in *R*_t_ over time reported for COVID-19 (Yang et al 2020). We therefore consider the impact of different values of the initial reproductive ratio (figure 4), which could capture this social aspect, or could represent heterogeneity between individuals in the amount of virus shed, or could inform about innate differences in behaviour between China and the UK. Given the strong biasing of transmission towards long-duration contacts, the impact of varying initial reproductive ratio is less extreme than might be expected; it is only for the highest values of initial reproductive ratio simulated (>9.8) that contact tracing fails to find more than one case such that infection can escape.

**FIGURE 4.**
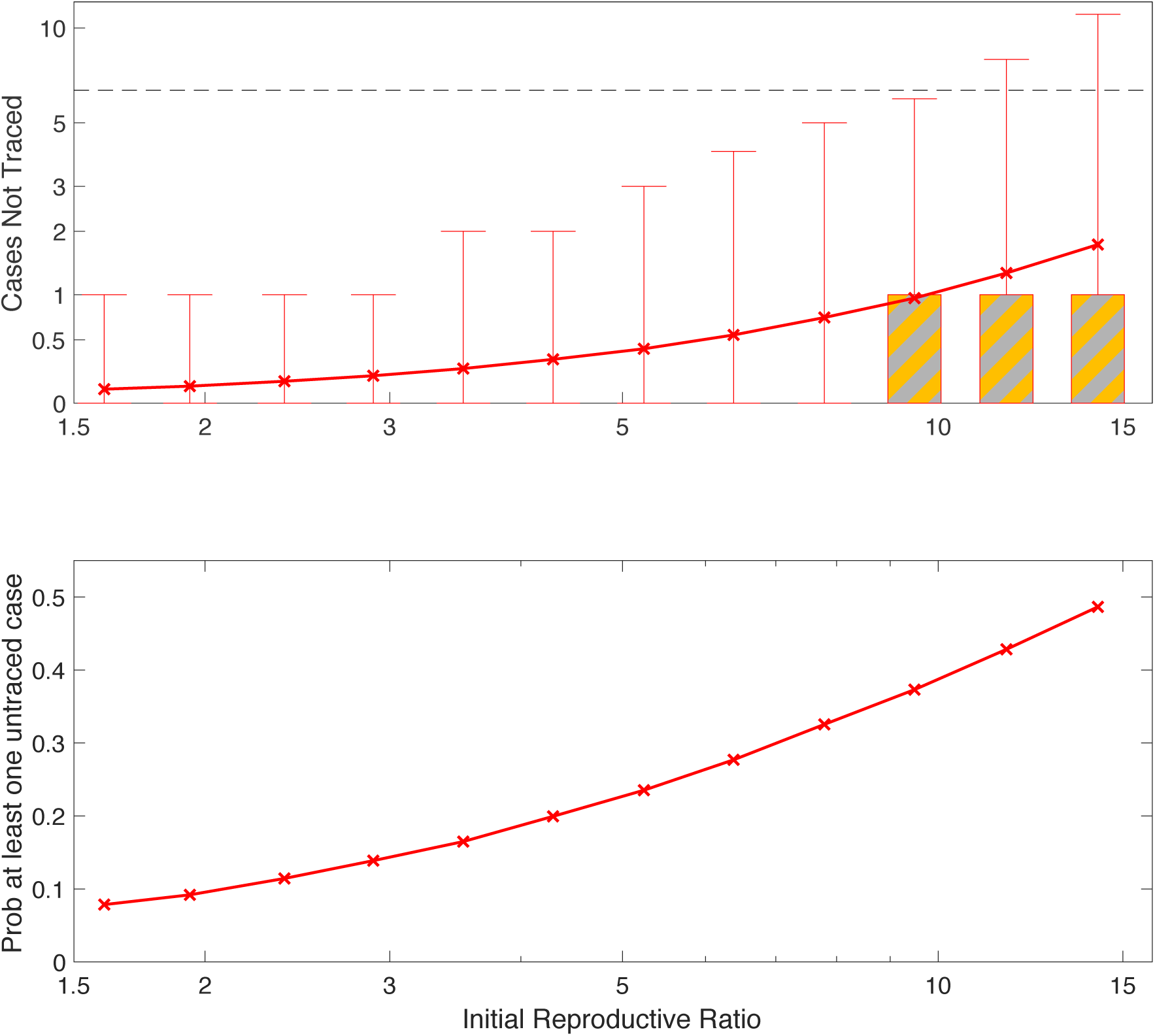
Impact of different values for the initial reproduction number of the primary case; changing this does not affect the the number of contacts traced (a) the number of secondary contacts that are not traced (b) the probability that at least one secondary case is not traced. For (a) the crosses mark the mean value, boxes contain the 50th percentiles while bars contact the 95th percentiles, colours correspond to those in Figure 1a - distributions are across all respondents to the survey and across stochastic realisations. (Based on an SEIR model with latent period 4, infectious period 1.61, *R*_0_=3.11).

## Conclusions

Mathematical models have an important role to play in preparedness for novel infectious diseases, allowing policy makers to plan for potential public health scenarios before they arise. However, in such scenarios reliable data is often limited, so predictions of long term dynamics are generally associated with wide confidence intervals. In contrast, while short term predictions are subject to greater stochasticity, the distribution of possible behaviours can be readily captured. Here we have investigated contact tracing of a close-contact pathogen, using 2019 novel coronavirus (COVID-19) as the example, and considered the efficacy of contact tracing as a control measure. This work brings together a detailed survey of social encounters together with bespoke mathematical modelling of the transmission and tracing processes. Given the huge heterogeneties present in social encounters (both in terms of duration and number) mathematical models are vital to interpret the interplay between a low number of high risk encounters (e.g., household members) and the high number of low-risk less-identifiable encounters (e.g., commuters or retail customers).

The UK currently defines a close contact as 15 minutes within 2 meters over two weeks before detection (PHE 2020). Under this definition, there are unlikely to be many untraced secondary cases, although the burden of tracing could be large. Relaxing the definition of a contact (such that longer contact durations are needed) lessens this burden but at the greater risk of undetected cases (Figure 3). Surprisingly, small changes to the reproductive ratio, within the bounds estimated from early data (Figure 4) or even changes to the distribution of infectivity, are predicted to have a relatively modest impact of the success of contact tracing illustrating the robustness of this control measure.

Our model has addressed the simple and optimistic question of whether contact tracing is sufficient to identify secondary infections. The public health implications of this tracing are more complex, and depend on the relative timing of events and the treatment of identified contacts. For contact tracing to be an effective public health measure requires secondary cases to be discovered before they become infectious; hence the time from the primary case becoming infectious to the tracing of their contacts needs to be shorter than the incubation period. Longer time scales would allow tertiary cases to be infected and would snowball the tracing process. In addition, those contacts that are traced either need to be effectively screened for infection and quarantined or otherwise isolated so that they do not pose a risk to others. Therefore, while contact tracing has the potential to control COVID-19 (and other close-contact pathogens) the ultimate success relies on the speed and efficacy with which suspect contacts can be contained.

## Data Availability

Social Contact Survey data is available from http://wrap.warwick.ac.uk/54273/

## APPENDIX

From the contact tracing data, we extrapolate to the estimate the duration of contact 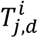 between individual *i* (the respondent) and individual *j* (the contact) on day *d*.

We then define a close contact of *i* as all contacts *j*:

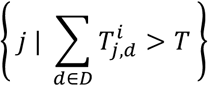

where in the UK, we have defined the total contact time *T* as 15 minutes over a duration *D* of two weeks before detection and isolation of individual *i* (PHE 2020).

The probability of transmission to individual *j* from individual *i* is then calculated as:

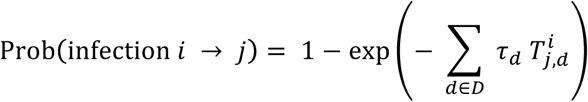

Where *τ*_*d*_ is an estimate of the transmission rate from individual *i* on day *d*.

